# Selective Bacteroidota Depletion but Not Global Microbiome Differences in Short Bowel Syndrome on Fiber-Free Enteral Nutrition

**DOI:** 10.64898/2026.10.27.26366621

**Authors:** Wenjing Zong, Scott G. Daniel, Christina M. Zarek, Naomi G. Wilson, Clarice Cook, Natalie Riebe, Jacqueline Kirsch, Elicenda Tovar Marquez, Reena Alame, Nandini Channabasappa, Van Nguyen, Kyle Bittinger, Gary D. Wu, Christina Bales, Andrew Y. Koh, Lindsey Albenberg

## Abstract

Intestinal dysbiosis in pediatric short bowel syndrome (SBS) is associated with poor growth, but the factors that are driving this dysbiosis remain unclear. Prior studies suggest that parenteral nutrition (PN) dependence and remnant intestinal anatomy are correlated with dysbiosis. However, these studies often compared SBS patients to healthy controls, making it difficult to distinguish SBS-specific factors from alterations due to differences in diet, namely the consumption of enteral nutrition with complex composition and exposure to dietary fiber. In this preliminary report from the GREENBEANS study, we examined baseline stool shotgun metagenomic profiles from 10 SBS patients and 13 non-SBS controls receiving fiber-free enteral nutrition. Alpha diversity did not differ significantly between groups by richness or Shannon diversity. Beta diversity showed minimal separation by Bray-Curtis distance and modest separation in Jaccard distance. Differential abundance analysis identified depletion of Bacteroidota in SBS. Compared to control patients also receiving fiber-free enteral nutrition, SBS was associated with only a small global difference in the gut microbiome, suggesting that diet, and not anatomy as previously assumed, is the predominant determinant of gut microbiome structure. This suggests that the inclusion of complex substrates, such as dietary fiber, in enteral nutrition, may be a modality to prevent the development of a dysbiotic microbiome in patients with SBS.

**What is known/What is new:** *What is known:*

- Children with short bowel syndrome (SBS) often demonstrate intestinal dysbiosis characterized by reduced microbial diversity and enrichment of Proteobacteria/Enterobacteriaceae compared with healthy controls.
- Dysbiosis in SBS is associated with increased PN dependence and poor growth outcomes.
- The relative contribution of various SBS-associated factors to this dysbiosis remains unclear because prior studies generally used healthy control subjects.

*What is new:*

- SBS-specific differences while on fiber-free enteral nutrition were driven primarily by selective depletion of Bacteroidota anaerobes rather than broad Proteobacteria expansion.
- SBS was associated with only a modest global microbiome separation from non-SBS controls when both were receiving fiber-free enteral nutrition, suggesting that the lack of dietary fiber, rather than the alteration of intestinal anatomy or other SBS-specific factors, may be a major factor in the development of the SBS-associated dysbiosis found in prior studies.
- Our findings shift attention from fixed anatomic factors alone towards potentially modifiable dietary contributors in SBS-associated dysbiosis, including enteral substrate complexity and dietary fiber exposure.

## Introduction

Short bowel syndrome (SBS) is the leading cause of pediatric intestinal failure^1^. SBS patients have reduced absorptive capacity that results in prolonged dependence on parenteral nutrition (PN) and require specialized enteral nutrition (EN)^1,2^. The gut microbiome has emerged as a potential modifier of intestinal adaptation and clinical outcomes in SBS. Prior pediatric studies have described marked dysbiosis in SBS, featuring reduced microbial diversity, enrichment of Proteobacteria or Enterobacteriaceae, and depletion of beneficial commensal anaerobes in association with increased PN dependence and growth outcomes^3-6^. These studies support the relevance of dysbiosis in important clinical outcomes in SBS.

What drives SBS-associated dysbiosis is unclear, however. Microbiome alterations in SBS may reflect altered anatomy, abnormal motility, PN dependence, specialized EN, or frequent antibiotic exposure. Children with SBS often receive specialized EN that is hydrolyzed (peptide-based) or elemental (amino acid-based)^2,7,8^. These formulas are often devoid of dietary fiber. Yet dietary fiber promotes diverse, healthy microbiota, as it is a fermentable nutrient for beneficial taxa^7,9-11^. Prior studies comparing SBS to healthy children on normal diets could not completely account for these extreme differences in nutritional composition and enteral substrate complexity^4,5^. To evaluate the SBS microbiome under a similar EN composition context, we compared baseline stool shotgun metagenomic profiles from children with SBS and non-SBS controls who were both receiving fiber-free EN.

## Methods

### Ethics Statement

Participants were enrolled in the GREENBEANS study (ClinicalTrials.gov NCT05432648), a prospective interventional study conducted at the Children’s Hospital of Philadelphia (CHOP) and the University of Texas Southwestern (UTSW) investigating the effects of fiber-food introduction (green bean puree) in children with short bowel syndrome (SBS) vs non-SBS controls. The study was approved by the respective Institutional Review Boards (IRB# 21-019185 and STU-2023-1152). Written informed consent and/or assent was obtained from participants or their legal guardians, as appropriate.

### Patient Population and Study Design

Eligible participants were 4 months adjusted age to 17 years. The present study represents an interval analysis of baseline samples and clinical metadata collected prior to any dietary fiber introduction, where both groups were receiving fiber-free EN. All SBS participants had a confirmed diagnosis of SBS based on surgical and/or imaging records, a history of PN dependence for greater than 2 months, a fiber-free EN tube feeding regimen and were followed in the Intestinal Rehabilitation Program of their respective institutions. SBS inclusion criteria included: less than 20% calories from oral foods, at least 20% calories from a tube feeding formula without fiber, remaining calories may be from EN and/or PN. A stable regimen of antibiotics starting from 2 weeks prior to intervention was allowed, to be inclusive of SBS patients consistently on antibiotics for small intestinal bacterial overgrowth (SIBO). Non-SBS controls were recruited from the outpatient Gastroenterology Clinic at CHOP and consisted of children without SBS who were receiving a fiber-free EN, with less than 20% calories from oral foods. The non-SBS control group did not have a unifying diagnosis; most had neurodevelopmental or feeding disorders requiring long-term tube feeding. Exclusion criteria for both groups included primary gastrointestinal dysmotility disorder, mucosal intestinal, malignancy, immunodeficiency, or new antibiotic exposure within 14 days of enrollment.

### Nutritional History

Nutritional data were abstracted from registered dietitian assessments performed within 3 months of biospecimen collection and supplemented by a NHANES-adapted standardized dietary frequency questionnaire^12^. Caloric intake from food included orally consumed foods, fiber-containing formulas, or food-based blenderized feeds (consumed orally or through tube feeding). Caloric intake from EN included fiber-free milk, including polymeric, hydrolyzed, elemental formulas, or breast milk. Caloric intake from PN consisted of hyperalimentation, intravenous fat emulsion, and any dextrose-containing fluids.

### Antibiotic History

Antibiotic exposure was categorized as current (active antibiotic use at the time of sample collection or completion of an antibiotic course within the preceding 14 days) or none (no antibiotic use within 14 days of sample collection).

### Sample Collection

Stool samples were obtained during a scheduled outpatient clinic visit or via a standardized home collection kit on enrollment. Stool samples were processed according to study protocols and stored at −80°C until analysis.

### Shotgun Metagenomic Sequencing and Analysis

Shotgun metagenomic sequencing was performed on stool sample DNA as previously described (Illumina platform)^10^. Sequence quality control, host read removal, and taxonomic classification of bacteria were performed using the Sunbeam pipeline with Kraken. Additional details regarding DNA extraction, library preparation, sequencing parameters, and bioinformatic processing are provided on request. Statistical analyses were performed using R (version 4.5.1).

Alpha diversity was assessed using observed species richness (rarefied to a sampling depth of 10,000 reads) and Shannon diversity index. Differences in alpha diversity between SBS and non-SBS groups were evaluated using linear models with SBS status as the primary predictor.

Beta diversity was assessed using Bray-Curtis dissimilarity and Jaccard distance and visualized by principal coordinates analysis (PCoA). Community-level separation between groups was evaluated by PERMANOVA using the adonisplus package, with SBS group membership permuted between subjects. The proportion of variance explained by SBS status (R^2^) is reported alongside p-values.

Differential abundance analysis was restricted to taxa with mean relative abundance greater than 1% across all samples. Relative abundances were log10-transformed, and differences between SBS and non-SBS groups were estimated by linear models with SBS status as the predictor. P-values were corrected for multiple comparisons using the Benjamini–Hochberg false discovery rate (FDR) method. FDR < 0.05 was considered statistically significant.

To evaluate robustness to potential confounders, sensitivity analyses were conducted with age and antibiotic use added as covariates, separately and in combination. For differential abundance, sensitivity models were applied to FDR-significant taxa.

Clinical data were collected and stored using REDCap, a secure, web-based application designed to support data capture for research studies.

## Results

### Patient characteristics

Baseline clinical and nutritional characteristics are summarized (**Table 1**). The cohort included 13 non-SBS controls and 10 children with SBS. Age and sex distribution were similar between groups, with a median age of 2.0 years [IQR 8.9] and 1.9 years [IQR 4.8], and female sex of 54% and 60%, in non-SBS controls and in children with SBS, respectively. Both groups were receiving fiber-free EN and/or PN, with minimal calories from fiber-foods, fiber-formula, or oral intake. However, fiber-free formula type differed between the two groups, with non-SBS controls receiving a variety of polymeric formulas or breast milk. All children with SBS received elemental formula, a common clinical practice^2^. Among children with SBS, the median residual small bowel length was 61.5 cm [IQR 46.3–82.0], corresponding to 39.5% [IQR 28.3–61.3] of expected length. Only 2 children with SBS had an ileocecal valve, and 6 had partial or minimal colon remaining. Four children with SBS were receiving antibiotics, most commonly metronidazole for empiric SIBO treatment, while no non-SBS controls were receiving antibiotics.

**Table 1.**
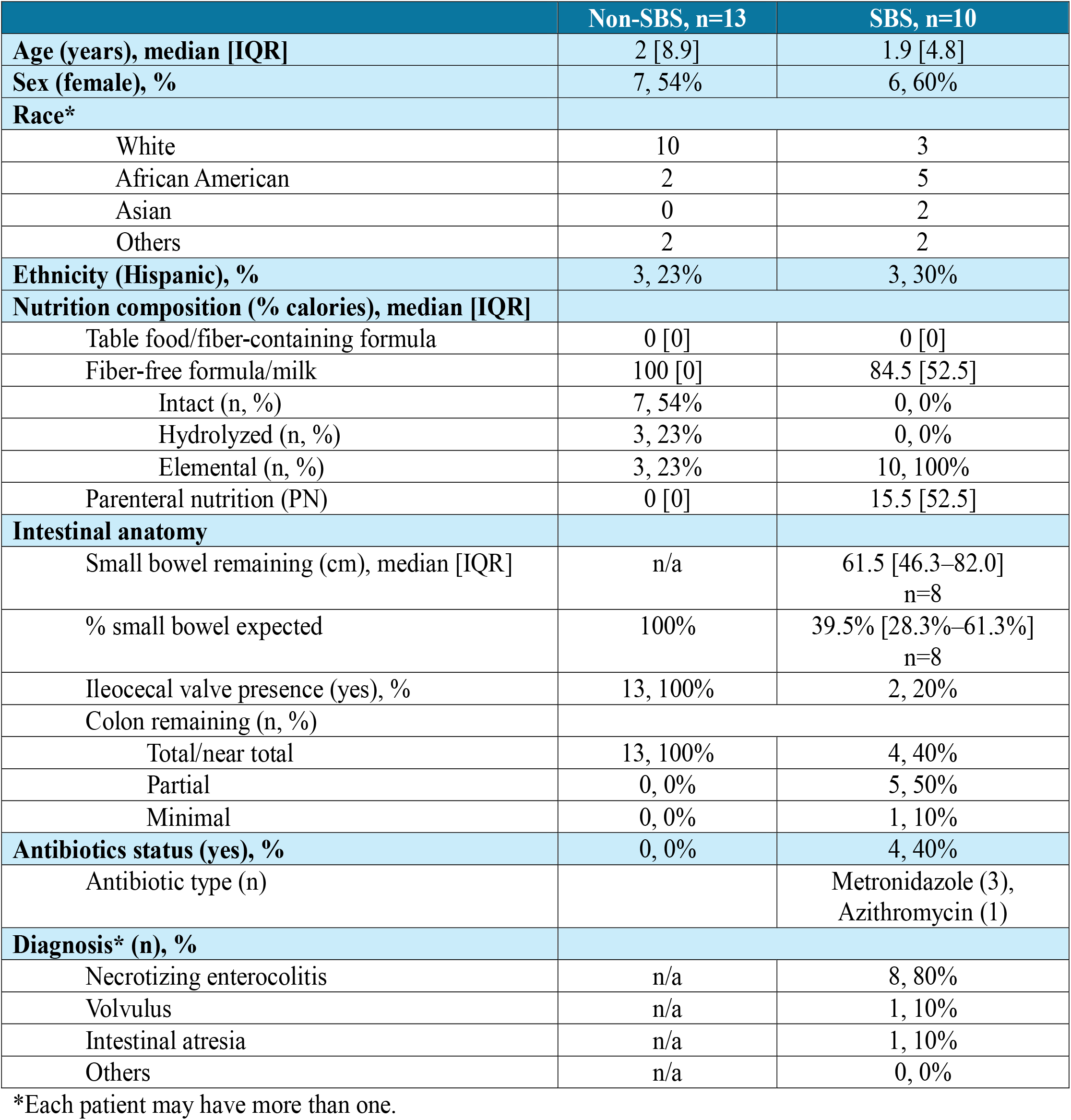
Clinical characteristics of non-SBS controls and SBS children. Values are median [IQR] or n, (%). Nutrition composition represents percentage of total caloric intake. Continuous variables were compared using Wilcoxon rank-sum test and categorical variables using Fisher’s exact test. SBS-specific anatomy variables were summarized among children with available operative documentation; missing values were not imputed.

### Microbiome structure in fiber-free states

Baseline stool shotgun metagenomic profiles were compared between non-SBS controls and SBS patients. We hypothesized that the marked differences in intestinal anatomy and antibiotic exposure would significantly alter microbial communities between controls and SBS children. Surprisingly, alpha diversity, a measure of bacterial diversity within an individual, was not significantly different in patients with SBS compared to non-SBS patients, both of which are consuming fiber-free EN (**Figure 1A**). This suggests that the altered anatomy in patients with SBS is not the primary driver in the marked reduction in overall richness or evenness (Shannon index) in this fiber-free setting.

**Figure 1.**
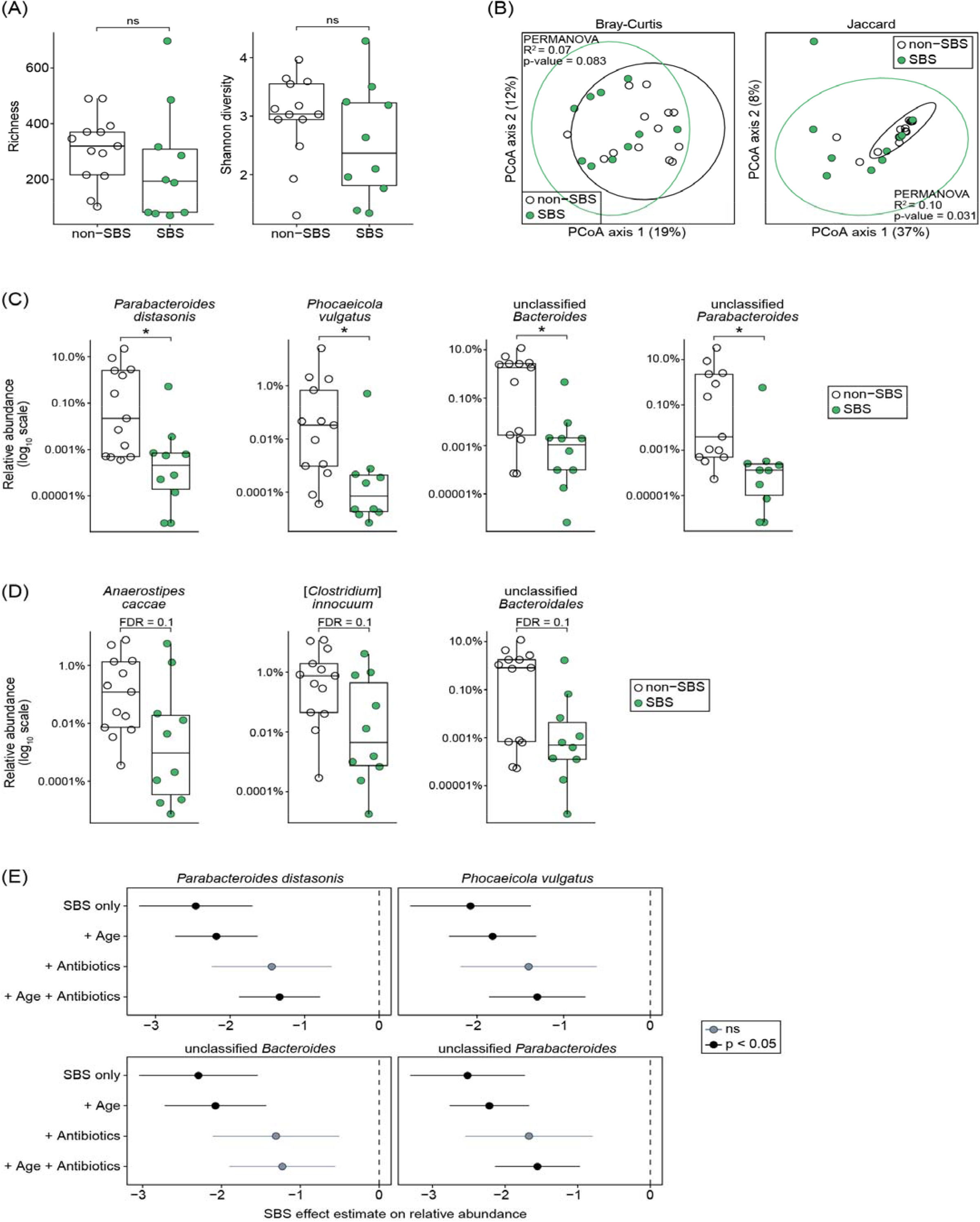
Gut microbiome diversity and community structure in children receiving fiber-free enteral nutrition. **A-D)** Baseline stool shotgun metagenomic profiles were compared between non-SBS and SBS children. **A)** Alpha diversity analysis (Richness and Shannon diversity). **B)** Beta diversity analysis (Bray-Curtis distance and Jaccard distance with 95% confidence ellipse) visualized by principal coordinates analysis. **C)** Relative abundance of significantly different taxa. **D)** Relative abundance of taxa with significant p-values, but insignificant FDR p-values, between non-SBS and SBS patients. **E)** Sensitivity analyses for four significantly altered Bacteroidota taxa (effect estimates on log_10_-transformed relative abundance). Negative effect estimates indicate lower relative abundance in SBS compared with non-SBS controls. Points indicate effect estimates and lines indicate standard errors. Each point = data from an individual stool sample (A-D); Boxes = median and IQR, whiskers = 1.5 x IQR (A, C, D). Linear models (A), PERMANOVA (B), Linear models with Benjamini-Hochberg false discovery rate correction (C-D). \**p* < 0.05, ns, not significant. SBS, short bowel syndrome; PCoA, Principal Coordinates Analysis; FDR, false discovery rate; IQR, interquartile range.

We next evaluated overall community composition using beta diversity analyses (**Figure 1B**). Bray-Curtis dissimilarity demonstrated minimal separation between groups, with SBS status explaining 7% of variation in Bray-Curtis distance (PERMANOVA, p=0.083), indicating substantial overlap in community structure and taxa abundances. Jaccard distance analysis showed greater separation, with SBS status explaining 10% of variation (PERMANOVA, p=0.031). These findings indicate partial separation in taxon membership, but substantial overlap in overall community structure between non-SBS controls and children with SBS when both are consuming fiber-free EN.

Although global diversity measures were similar, differential abundance analysis identified selective taxonomic differences between groups. Children with SBS had lower relative abundance of several Bacteroidota taxa (**Figure 1C**, FDR=0.045), including *Parabacteroides distasonis, Phocaeicola vulgatus* (formerly *Bacteroides vulgatus*), unclassified *Bacteroides*, and unclassified *Parabacteroides*. These taxa are associated with dietary fiber metabolism^13,14^. Additional taxa, including *Anaerostipes caccae, Clostridium innocuum*, and unclassified *Bacteroidales*, were nominally different between groups but did not meet the false discovery threshold (**Figure 1D**). In sensitivity analyses, the SBS-associated differences in *P. distasonis* and *P. vulgatus* remained significant after adjustment for age, whereas adjustment for antibiotics attenuated the effect estimates (**Figure 1E**). These findings suggest that Bacteroidota depletion in SBS is not explained by age but may be partly influenced by antibiotics. Contrary to the expectation that SBS-associated dysbiosis would be dominated by expansion of Proteobacteria/Enterobacteriaceae as previously described^3^, group differences in the fiber-free setting were likely driven by decreases in Bacteroidota.

## Discussion

The current study was designed to reduce nutritional composition confounding by comparing children with SBS to non-SBS controls who were both receiving fiber-free diets through tube-feeds or by mouth. In this interval analysis of an ongoing clinical trial, children with SBS demonstrated minimal microbiota community separation from non-SBS controls despite substantial differences in intestinal anatomy, PN exposure, formula type, and antibiotic exposure. Alpha diversity did not differ significantly between groups, and beta diversity analyses showed overlapping community structure. However, SBS was associated with selective depletion of Bacteroidota anaerobes, including *Parabacteroides distasonis* and *Phocaeicola vulgatus*. Notably, SBS was not dominated by broad Proteobacteria expansion, historically associated with SBS-dysbiosis^3^. This likely reflects our unique study design, which controls for the lack of dietary fiber. In our model, SBS associated dysbiosis is likely driven by reduced luminal nutrient delivery and greater reliance on fiber-free, elemental formula.

Enteral nutrition is widely considered the major driver of intestinal adaptation, providing luminal stimulation that promotes epithelial growth, gastrointestinal hormone secretion, and pancreaticobiliary stimulation^15^. SBS nutrition reviews and guidelines emphasize progressive advancement of enteral or oral nutrition while reducing PN^2^. Importantly, higher nutrient complexity has been proposed to provide greater digestive workload and luminal stimulation, potentially supporting adaptation^15^. Our findings extend this concept to the microbiome. When enteral substrate is low in diversity, such as in fiber-free states, the microbiota may be constrained regardless of SBS status. Conversely, increasing enteral substrate complexity (namely, the addition of fermentable dietary fiber), when clinically safe, may represent a microbiome-directed strategy towards intestinal rehabilitation. Fermentable fiber may be particularly relevant in SBS because colonic bacterial fermentation of unabsorbed carbohydrates generates short chain fatty acids (SCFA), which can provide additional energy salvage and support intestinal adaptation^7,11,15^.

The selective depletion of Bacteroidota taxa in SBS is biologically plausible. Members of Bacteroidota are prominent anaerobic gut commensals with capacity for complex carbohydrate and glycan metabolism^13^. Their lower abundance in SBS may reflect reduced availability of complex enteral substrates (elemental vs intact formula/breast milk), faster transit, altered bile acid metabolism, and/or antibiotic exposure. This finding is also consistent with our prior work, where children with SBS had reduced Bacteroidetes compared with healthy controls, and greater whole-foods intake within SBS was associated with higher Bacteroides abundance^8^. Antibiotic exposure is particularly relevant in this cohort because 40% of our SBS participants were receiving antibiotics, while no control patients received antibiotics. The most common antibiotic was metronidazole, used for SIBO, which may suppress anaerobic taxa. The attenuation of the *P. distasonis* and *P. vulgatus* signal after antibiotic adjustment suggests that cycling antibiotics may also contribute to the SBS-associated depletion of these anaerobic taxa.

This study has several limitations. First, the sample size was small. Additionally, while SBS and non-SBS groups were both on fiber-free EN, they differed in formula type, PN exposure, and antibiotics exposure. These factors are clinically inseparable from SBS severity and limit attribution of microbiome differences to intestinal anatomy alone. In addition, stool samples do not fully represent small intestinal microbial communities, which may be particularly relevant in SBS.

Our findings have clinical implications. Intestinal dysbiosis is associated with growth outcomes in SBS, yet it is not easily modifiable by interventions such as probiotics^3,16^. This study challenges the assumption that SBS anatomy alone explains the marked dysbiosis described in prior cohorts and raises nutrition composition (specifically the lack of dietary fiber) as an important contributor. This distinction has important implications. If the dysbiosis is driven primarily by fixed anatomic factors associated with SBS diagnosis, it may be difficult to modify. However, if part of the dysbiosis reflects restricted EN complexity, then gradual expansion of EN complexity, including fiber-containing foods or formulas when clinically appropriate, represents an actionable approach within intestinal rehabilitation. Our findings support future prospective studies testing whether fiber or whole-food introduction can modify microbiome structure and improve clinically meaningful outcomes such as growth and PN weaning.

## Conclusion

In conclusion, children with SBS showed only modest global microbiome separation from non-SBS controls when both receive fiber-free diets but demonstrated selective depletion of Bacteroidota anaerobes. These findings suggest that reduced enteral substrate complexity may be an important ecological constraint on gut microbiome structure in children regardless of diagnosis, while SBS-specific factors such as anatomy, motility, PN dependence, and antibiotic exposure may selectively affect anaerobic taxa. Future studies should evaluate whether safe, gradual expansion of enteral substrate complexity, such as the introduction of fiber-containing foods and/or fiber supplements, can modify microbiome structure and function to improve clinical outcomes during pediatric intestinal rehabilitation.

## Data Availability

All data produced in the present study are available upon reasonable request to the authors

## Acknowledgements

We thank the Gastrointestinal Epithelium Modeling (GEM) Program at Children’s Hospital of Philadelphia for sample storage. We are grateful to the physicians, dieticians, and nurses at the Intestinal Rehabilitation Program at Children’s Hospital of Philadelphia and University of Texas Southwestern Medical Center.

